# Screening for Atrial Fibrillation in Older Adults at Primary Care Visits: the VITAL-AF Randomized Controlled Trial

**DOI:** 10.1101/2021.08.13.21261969

**Authors:** Steven A. Lubitz, Steven J. Atlas, Jeffrey M. Ashburner, Ana T. Trisini Lipsanopoulos, Leila H. Borowsky, Wyliena Guan, Shaan Khurshid, Patrick T. Ellinor, Yuchiao Chang, David D. McManus, Daniel E. Singer

## Abstract

**Background:** Undiagnosed atrial fibrillation (AF) may cause preventable strokes. Guidelines differ regarding AF screening recommendations. We tested whether point-of-care screening with a handheld single lead electrocardiogram (ECG) at primary care practice visits increases diagnoses of AF.

**Methods:** We randomized 16 primary care clinics 1:1 to AF screening using a handheld single-lead ECG (AliveCor KardiaMobile) during vital sign assessments, or usual care. Patients included were aged ≥ 65 years. Screening results were provided to primary care clinicians at the encounter. All confirmatory diagnostic testing and treatment decisions were made by the primary care clinician. New AF diagnoses over one-year follow-up were ascertained electronically and manually adjudicated. Proportions and incidence rates were calculated. Effect heterogeneity was assessed.

**Results:** Of 30,715 patients without prevalent AF (n=15,393 screening [91% screened], n=15,322 control), 1.72% of individuals in the screening group had new AF diagnosed at one year versus 1.59% in the control group (risk difference [RD] 0.13%, 95% confidence interval [CI] −0.16–0.42, P=0.38). New AF diagnoses in the screening and control groups differed by age with the greatest effect observed for those aged ≥ 85 years (5.56% versus 3.76%, respectively, RD 1.80%, 95% CI 0.18–3.30). The difference in newly diagnosed AF between the screening period and the prior year was marginally greater in the screening versus control group (0.32% versus −0.12%, RD 0.43%, 95% CI −0.01–0.84). The proportion of individuals with newly diagnosed AF who were initiated on oral anticoagulants was similar in the screening (n=194, 73.5%) and control (n=172, 70.8%) arms (RD 2.7%, 95% CI −5.5–10.4).

**Conclusions:** Screening for AF using a single-lead ECG at primary care visits was not associated with a significant increase in new AF diagnoses among individuals aged 65 years or older compared to usual care. However, screening may be associated with an increased likelihood of diagnosing AF among individuals aged 85 years or older and warrants further evaluation.

This study is registered with ClinicalTrials.gov, NCT03515057

**Funding:** Bristol Myers Squibb-Pfizer Alliance

## INTRODUCTION

Atrial fibrillation (AF) is a leading cause of stroke.^1^ Oral anticoagulation is effective for preventing strokes in patients with AF.^2^ However, AF may be asymptomatic and first diagnosed at the time of stroke.^3-5^ Screening may enable earlier diagnosis of AF and implementation of oral anticoagulation to prevent strokes.

Prospective uncontrolled studies have indicated that point-of-care screening for previously undiagnosed AF is feasible.^6-12^ Novel single-lead handheld electrocardiograms (ECGs) can enable rapid and scalable mass screening.^7,9-11^ Limited randomized controlled trial data addressing the effectiveness of point-of-care screening for detecting undiagnosed AF have produced varying results.^13-15^ Existing practice guidelines offer conflicting recommendations for AF screening. For example, the European Society of Cardiology (ESC) recommends opportunistic screening with either pulse palpation or ECG rhythm strip at clinic visits in patients at least 65 years of age.^16^ Similar guidelines from the National Heart Foundation of Australia and the Cardiac Society of Australia and New Zeland exist.^17^ The recommendations are in part predicated on randomized controlled trial data from the United Kingdom, performed in 2001 to 2003,^13^ which suggested that screening leads to a higher rate of AF detection. In contrast, the United States Preventive Services Task Force has stated that evidence is insufficient to recommend screening using ECGs.^18^

We aimed to assess whether routine screening of older adults using single-lead ECGs is more effective for diagnosing AF than usual care in a contemporary primary care practice setting. We hypothesized that such screening would identify more patients with AF, to enable efficient initiation of oral anticoagulation for those with confirmed AF at elevated stroke risk.

## METHODS

### Study design

The study methodology has been described previously.^19^ This was a pragmatic cluster-randomized controlled trial. Participating clinics were the unit of randomization. The research protocol was approved by the Mass General Brigham Institutional Review Board. Participants provided informed consent to participate. The study was considered minimal risk and a waiver of documentation of informed consent was granted. The conduct of the trial was monitored routinely by the investigative team.

### Participants

Sixteen of 22 practices within the Massachusetts General Hospital (MGH) Primary Care Practice Based Research Network were included and agreed to participate. Individuals aged at least 65 years presenting to a primary care clinician at a participating practice were included in the study population. Screening and control practices were initiated in a staggered manner at approximately one-month intervals, such that individuals were eligible to participate for a 12-month period in each practice. For simplicity, all patients, regardless of prior history of AF, were offered screening at the intervention practices. The impact of screening in patients with a prior diagnosis of AF will be reported separately. Encounters at which patients were not scheduled to see a primary care clinician (e.g., vaccinations) were not considered eligible study visits.

### Randomization

Cluster randomization at the practice level was used to facilitate implementation of the screening intervention and to minimize contamination in control patients. We used a constrained cluster randomization approach^20^ to optimize balance between the screening and control practices across factors that might influence the primary and secondary study endpoints among those without prior AF (**Supplemental Material**). We randomly assigned one of the two optimal paired groups to the screening intervention.

### Procedures

At all eligible visits during routine intake and vital signs assessment, clinic medical assistants asked eligible patients if they would like to participate in the study and briefly described the screening process. Consenting patients placed their fingers on a single-lead AliveCor KardiaMobile ECG device (AliveCor Inc., Mountain View, CA) affixed to an iPad (Apple Inc. Cupertino, CA) using the KardiaAI version 1 algorithm to conduct AF screening. Screening result categories included “Possible AF,” “Normal,” “Unclassified,” “No analysis (unreadable),” and “Patient declined screening.” Medical assistants were instructed to document the final screening results in the electronic medical record (Epic, Verona, WI) along with other vital signs and to notify primary care clinicians verbally if a patient had a “Possible AF” reading. All subsequent clinical management was determined by primary care clinicians, including follow-up 12-lead ECGs. Independent cardiologists reviewed all tracings within seven days and notified primary care clinicians if a pre-specified actionable rhythm was identified. Primary care clinicians and medical assistants were trained on study procedures at each screening practice before screening began and medical assistants had monthly refreshers.

### Outcomes

The primary outcome was incidence of newly diagnosed AF during the 12-month screening period. Secondary outcomes included change in the incidence proportion of AF from the 12-month window prior to the screening period (to account for possible differences in unscreened AF incidence rates in the two study arms), newly diagnosed AF associated with a primary care clinic visit, and a new medication list entry for an oral anticoagulant during the 12-month period after initial enrollment.

Newly diagnosed AF was initially identified from the electronic medical record.^21^ Individuals with an International Classification of Diseases, 10 Revision (ICD−10) code for AF or atrial flutter or a 12-lead electrocardiogram with AF or flutter in the diagnostic statement during the study period were identified. This was followed by adjudication of potentially new AF events as incident, prevalent,^22^ or not AF by a clinical endpoint committee comprising two research nurses and a cardiologist unaffiliated with the study and was based on a new medical record diagnosis of AF. The single-lead ECG tracing readings were not included in the adjudication of new AF. Newly diagnosed AF from the year prior to the study period was similarly ascertained and manually adjudicated. Further details of AF adjudication are provided in the **Supplemental Materials**.

Oral anticoagulation prescriptions were ascertained electronically using the electronic medical record. Oral anticoagulants included apixaban, dabigatran, edoxaban, rivaroxaban, and warfarin.

### Statistical analysis

Three patient samples were established for the analyses: (1) Intention-to-treat (group assignment was based on the patients’ first visit to a study practice during the study period); (2) per protocol (patients with a first visit to an intervention practice and screened at least once during the study period were assigned to the screening group and patients with first visit to a control practice and never screened were assigned to the control group); and (3) as treated (patients who were screened at least once during the study period were assigned to the screening group and patients never screened were assigned to the control group). Primary analyses were conducted using the intention-to-treat sample and sensitivity analyses were conducted using the “per protocol” and “as treated” samples.

For the primary outcome of AF incidence, unadjusted and adjusted generalized linear regression models that included established AF risk factors were used to compare the 2 groups. Risk differences (RDs) were estimated using Poisson models with identity link functions.^23^ Incidence rates per 100 person-years were compared between the two groups to account for varying length of follow-up among subjects. Exploratory analyses were used to investigate the heterogeneity of treatment effect by pre-specified factors (age, sex, heart rate, predicted risk of AF,^24^ implanted cardiac device, history of 12-lead ECG use, and number of primary care clinician visits in the previous year); no adjustment for multiple comparisons was applied due to the exploratory nature of the analyses. For the secondary outcome of change in AF incidence proportion from the 12-month window prior to the screening period, we included a time by group interaction in the models. Since outcomes obtained from patients cared for by the same clinician were not expected to be entirely independent, generalized estimating equations techniques were used to account for the clustering of patients within clinician data structure in all analyses.

The likelihood of incident AF detection on the same day of a primary care clinic visit in a study practice was compared between the two groups using a Poisson regression model with the generalized estimating equations approach to account for the repeated measures from the same individuals. We compared the proportion of individuals with new oral anticoagulant prescriptions using the same approach. Statistical significance was defined as a 2-tailed P value ≤ 0.05 and all analyses were conducted using SAS version 9.4 (The SAS Institute, Cary, NC).

The study was designed to provide sufficient statistical power to address the primary outcome (difference in AF incidence proportion). Based on data over a one-year period in 2017, the sample size was estimated to be 14,159 per group, and an effective sample size of 12,845 per group after accounting for clustering of patients within clinicians. The study had 80% power to detect a 0.48% increase in AF incidence rate (1.60% to 2.08%) when 85% of the screening group subjects were screened.

## RESULTS

Between July 31, 2018 and October 8, 2019, 15,393 individuals in the screening and 15,322 in the control arm without prevalent AF had at least one eligible visit at a study practice during the one-year study period, corresponding to 38,880 and 40,450 encounters, respectively. The median number of visits per person was 2 (Inter-quartiles [IQs]: 1–3). Overall, patient features in the screening and control arms were well-balanced. Characteristics of the practices and patients participating in the trial are provided in **Table 1**.

**Table 1.**
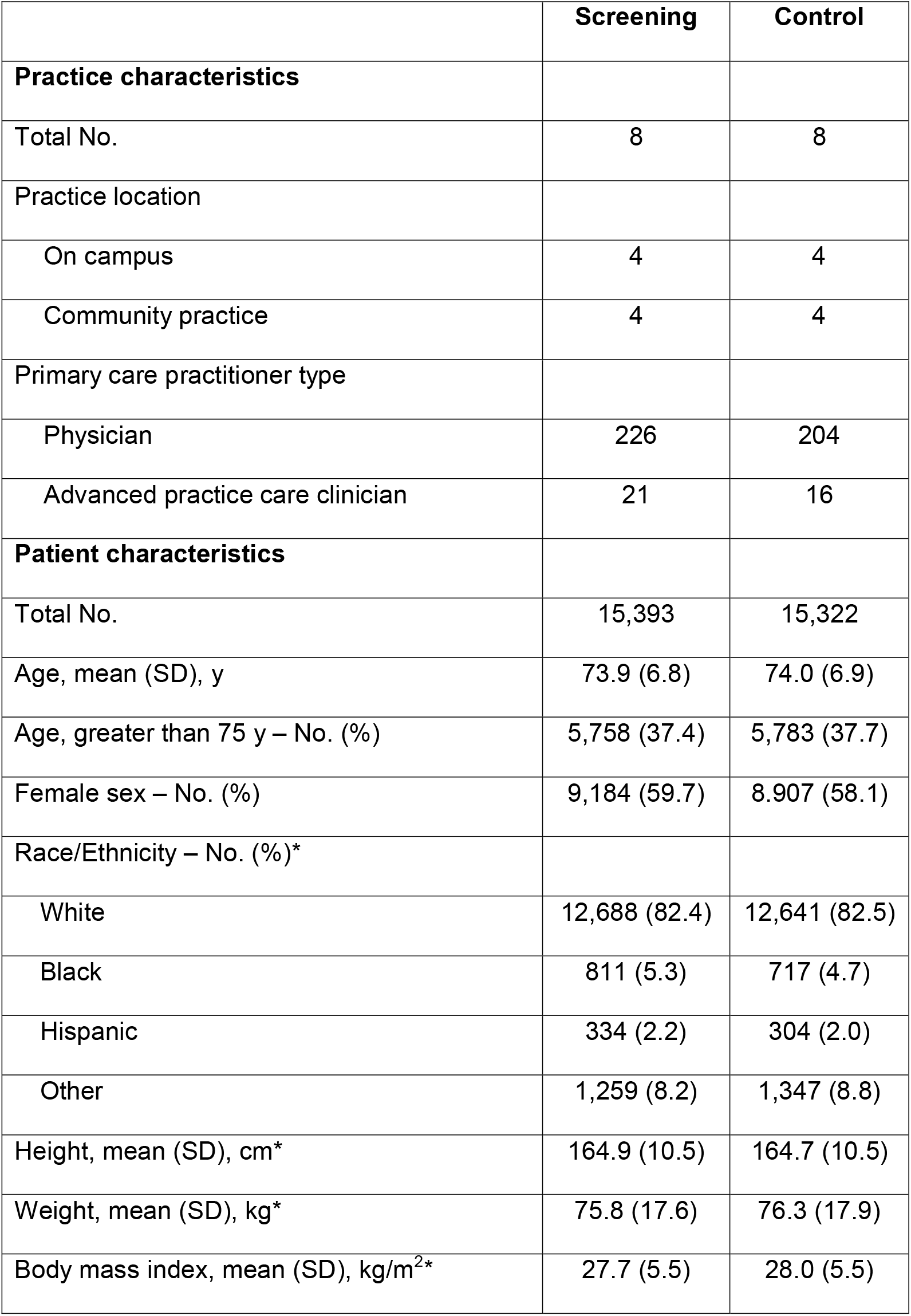
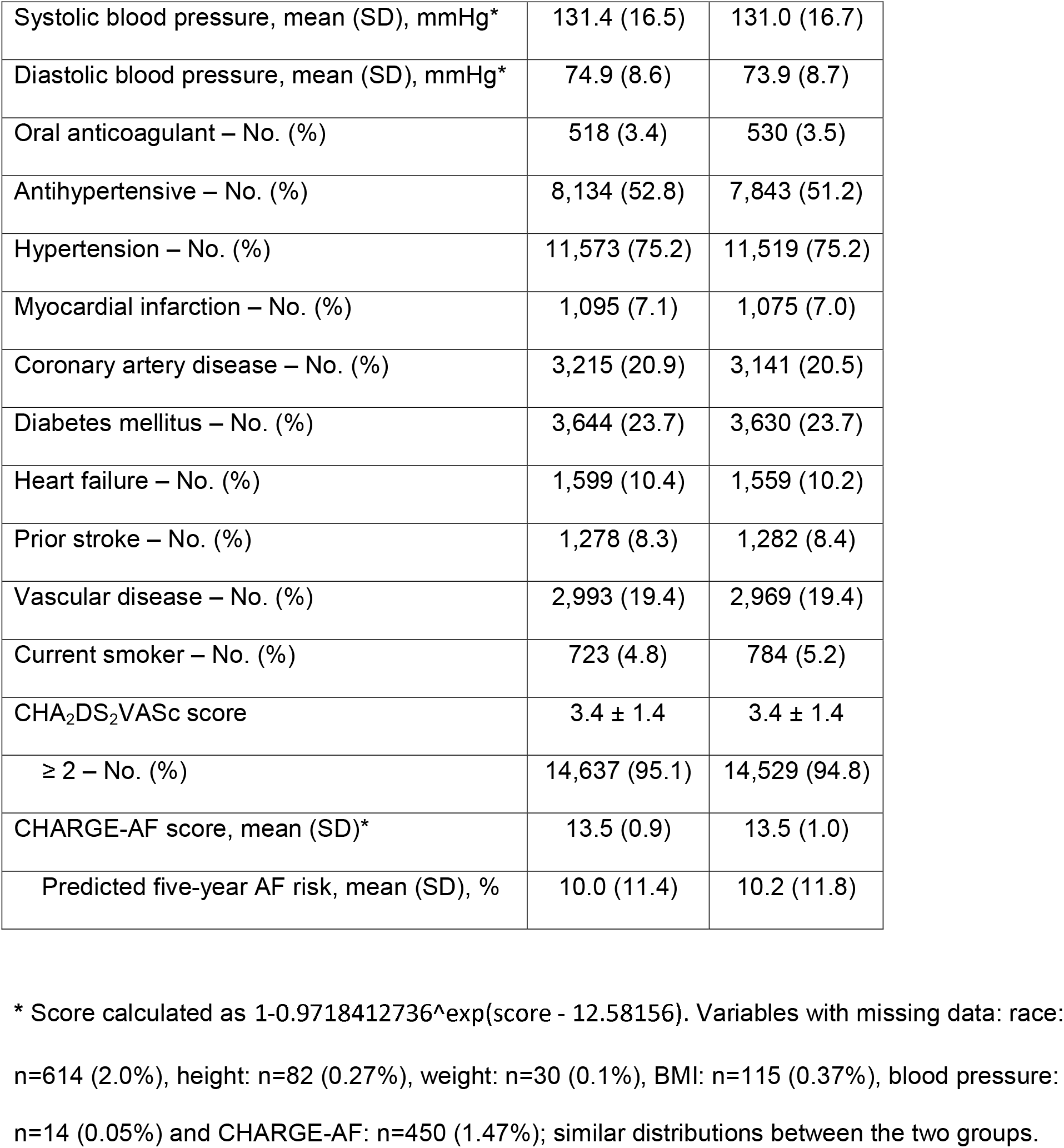
Practice and participant characteristics among individuals without prevalent atrial fibrillation.

|  | Screening | Control |
| --- | --- | --- |
| **Practice characteristics** |  |  |
| Total No. | 8 | 8 |
| Practice location |  |  |
| On campus | 4 | 4 |
| Community practice | 4 | 4 |
| Primary care practitioner type |  |  |
| Physician | 226 | 204 |
| Advanced practice care clinician | 21 | 16 |
| **Patient characteristics** |  |  |
| Total No. | 15,393 | 15,322 |
| Age, mean (SD), y | 73.9 (6.8) | 74.0 (6.9) |
| Age, greater than 75 y – No. (%) | 5,758 (37.4) | 5,783 (37.7) |
| Female sex – No. (%) | 9,184 (59.7) | 8,907 (58.1) |
| Race/Ethnicity – No. (%)* |  |  |
| White | 12,688 (82.4) | 12,641 (82.5) |
| Black | 811 (5.3) | 717 (4.7) |
| Hispanic | 334 (2.2) | 304 (2.0) |
| Other | 1,259 (8.2) | 1,347 (8.8) |
| Height, mean (SD), cm* | 164.9 (10.5) | 164.7 (10.5) |
| Weight, mean (SD), kg* | 75.8 (17.6) | 76.3 (17.9) |
| Body mass index, mean (SD), kg/m<sup>2</sup>* | 27.7 (5.5) | 28.0 (5.5) |
| Systolic blood pressure, mean (SD), mmHg* | 131.4 (16.5) | 131.0 (16.7) |
| Diastolic blood pressure, mean (SD), mmHg* | 74.9 (8.6) | 73.9 (8.7) |
| Oral anticoagulant – No. (%) | 518 (3.4) | 530 (3.5) |
| Antihypertensive – No. (%) | 8,134 (52.8) | 7,843 (51.2) |
| Hypertension – No. (%) | 11,573 (75.2) | 11,519 (75.2) |
| Myocardial infarction – No. (%) | 1,095 (7.1) | 1,075 (7.0) |
| Coronary artery disease – No. (%) | 3,215 (20.9) | 3,141 (20.5) |
| Diabetes mellitus – No. (%) | 3,644 (23.7) | 3,630 (23.7) |
| Heart failure – No. (%) | 1,599 (10.4) | 1,559 (10.2) |
| Prior stroke – No. (%) | 1,278 (8.3) | 1,282 (8.4) |
| Vascular disease – No. (%) | 2,993 (19.4) | 2,969 (19.4) |
| Current smoker – No. (%) | 723 (4.8) | 784 (5.2) |
| CHA<sub>2</sub>DS<sub>2</sub>VASc score | 3.4 ± 1.4 | 3.4 ± 1.4 |
| ≥ 2 – No. (%) | 14,637 (95.1) | 14,529 (94.8) |
| CHARGE-AF score, mean (SD)* | 13.5 (0.9) | 13.5 (1.0) |
| Predicted five-year AF risk, mean (SD), % | 10.0 (11.4) | 10.2 (11.8) |

\* Score calculated as $1 - 0.9718412736^{\exp(\text{score} - 12.58156)}$ . Variables with missing data: race: n=614 (2.0%), height: n=82 (0.27%), weight: n=30 (0.1%), BMI: n=115 (0.37%), blood pressure: n=14 (0.05%) and CHARGE-AF: n=450 (1.47%); similar distributions between the two groups.

Among individuals in the screening arm with at least one eligible practice visit during the study period, 14,047 (91%) underwent screening (**Figure 1**). Patients were approached for screening at 34,138 (89%) of the 38,502 eligible visits to intervention practices, and screening was conducted at 29,952 (78%) of eligible visits (median number of visits at which screening was conducted 2; IQ: 1–3).

**Figure 1.**
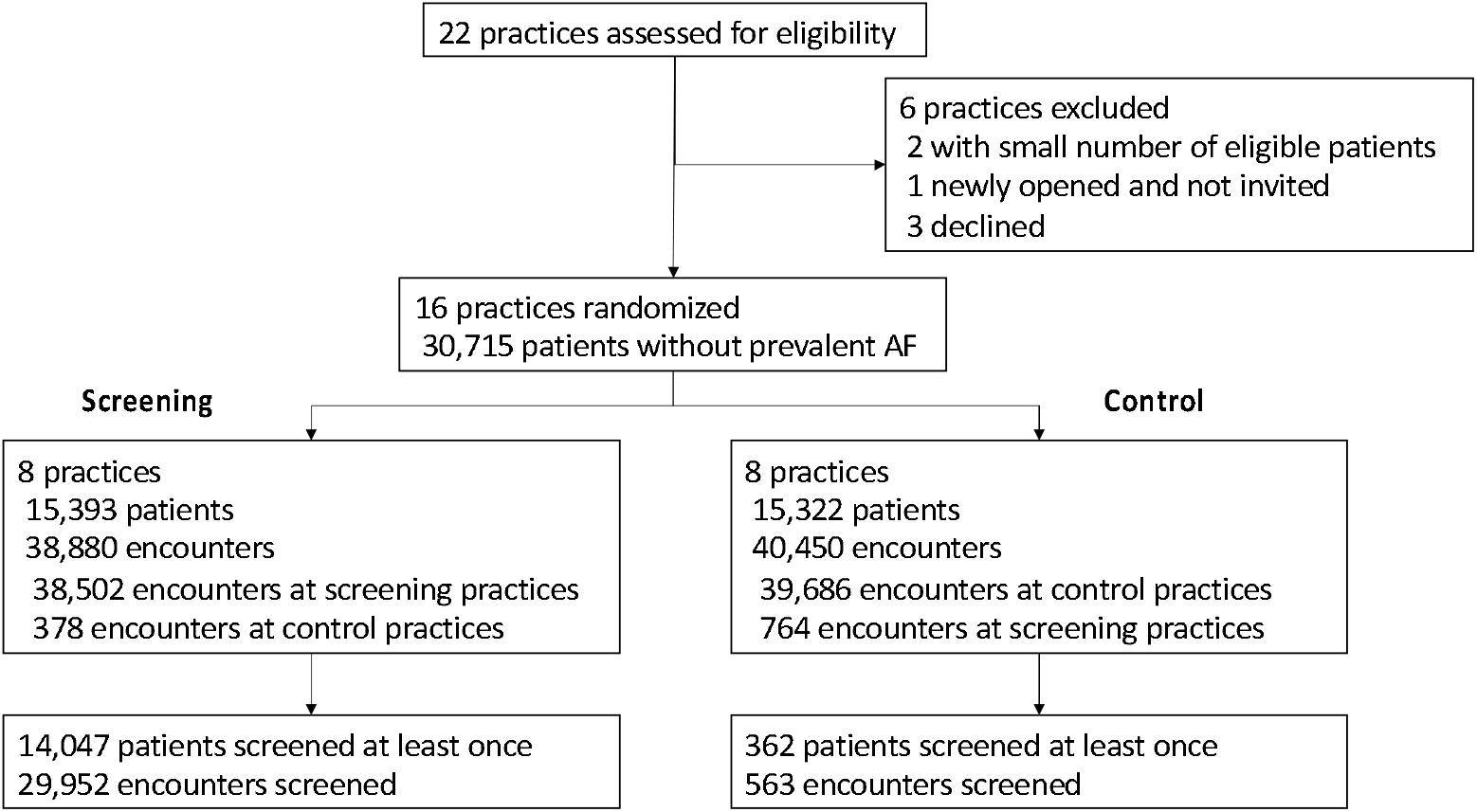
Practice and patient flow.

Among the 29,952 encounters to screening practices during which screening was conducted, the final automated single-lead ECG result was Normal (82.4%), Possible AF (3.0%), Unclassified (12.1%), or No Analysis (2.6%). In total, 5% of individuals screened had at least one automated single-lead ECG result of Possible AF. Same day 12-lead ECGs were ordered at encounters more commonly following an automated reading of Possible AF (46.6%) than encounters with Unclassified (9.9%), Normal (5.3%), and No Analysis (7.5%).

The primary outcome of newly diagnosed AF occurred in 264 (1.72%) individuals in the screening arm versus 243 (1.59%) in the control arm at one year (risk difference [RD] 0.13%, 95% CI −0.16–0.42, P=0.38, **Figure 2**). Ninety-six (36.4%) individuals with newly diagnosed AF in the screening arm had an AliveCor automated reading of Possible AF on or before the documented AF diagnosis during the study period. The AF incidence rate was 2.56 per 100 person-years for the screening group and 2.34 for the control group. Sensitivity analyses using the per protocol sample and the as treated sample showed similar results (**Supplemental Table 1**). Multivariable analyses adjusted for known predictors of AF also showed similar results (**Supplemental Table 2**).

**Figure 2.**
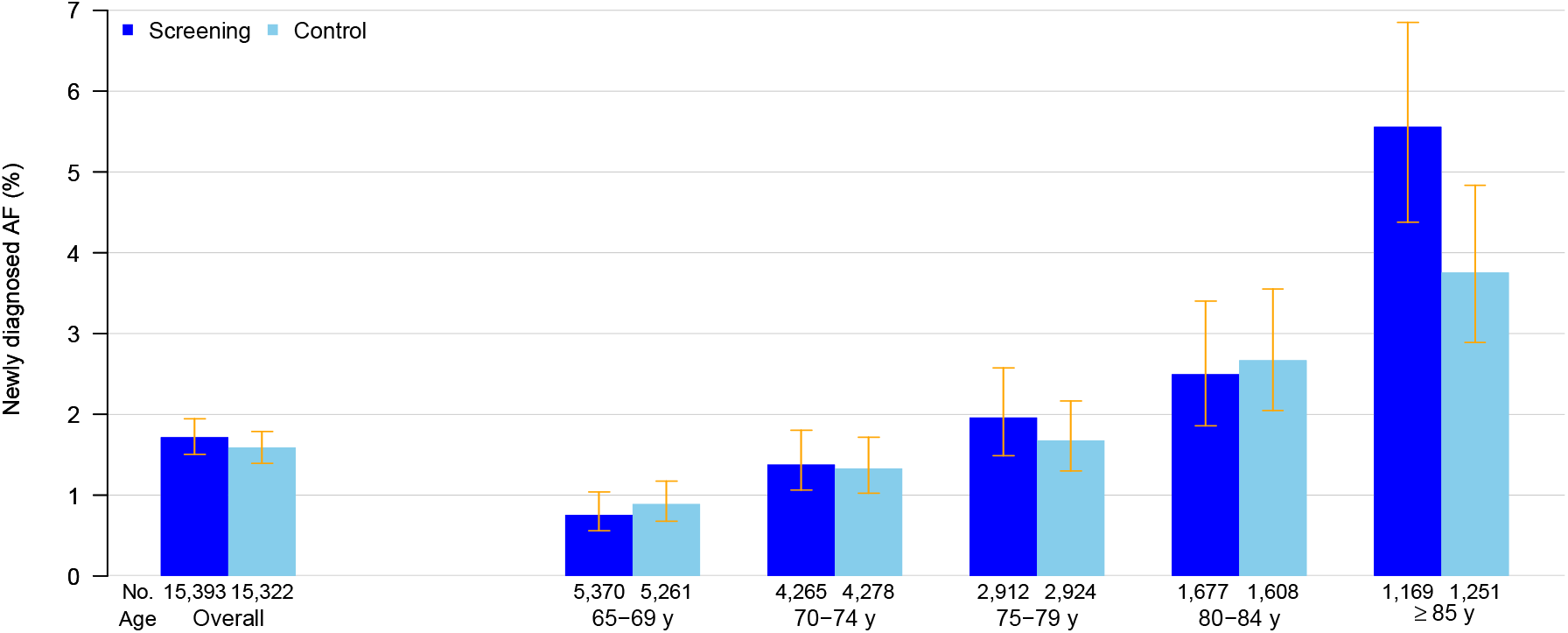
Proportion of individuals with newly diagnosed AF within 12 months in the screening and control groups overall and stratified by age.

The difference in proportion with newly diagnosed AF differed by patient age (**Figure 2**). Among individuals aged at least 85 years, new AF was diagnosed in 65 individuals (5.56%) in the screening arm versus 47 individuals (3.76%) in the control arm (RD 1.80%, 95% CI 0.18–3.30). There was little effect of the intervention across other subgroups (**Figure 3**).

**Figure 3.**
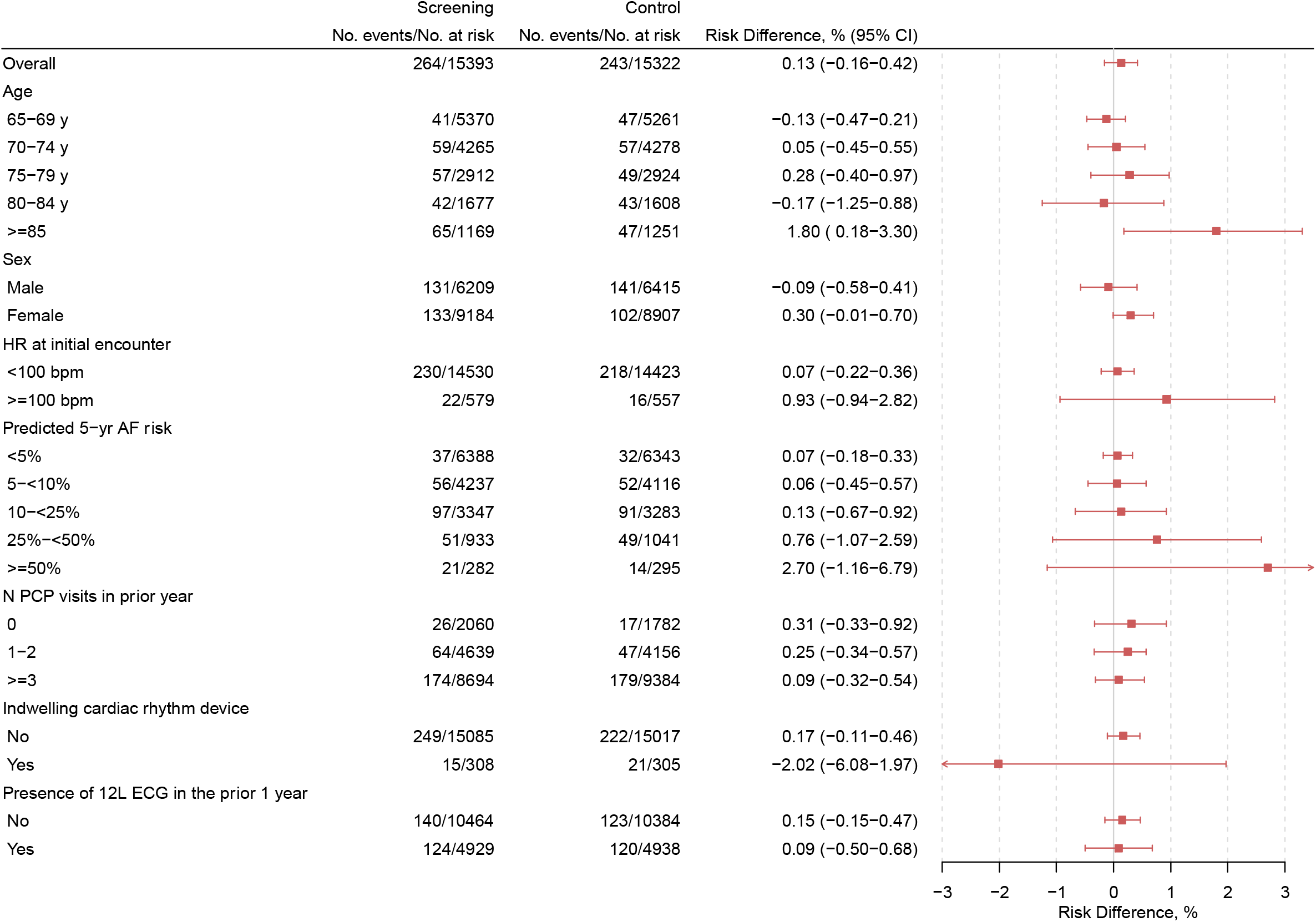
Subgroup analyses of screening for newly detected AF.

The difference in newly diagnosed AF between the screening period and the prior year was marginally greater in the screening versus control group (0.32% versus −0.12%, RD 0.43%, 95% CI −0.01–0.84). This effect of screening was primarily observed in patients 85 years and older (**Table 2**).

**Table 2.** Newly diagnosed atrial fibrillation in the year prior to versus the year of the screening intervention.

|  | Screening |  |  |  |  | Control |  |  |  |  | Difference in risk difference, % (95% CI) |
| --- | --- | --- | --- | --- | --- | --- | --- | --- | --- | --- | --- |
|  | Prior year |  | Screening period |  |  | Prior year |  | Screening period |  |  |  |
| Age category | N events / N at risk | % | N events / N at risk | % | Risk difference, % (95% CI) | N events / N at risk | % | N events / N at risk | % | Risk difference, % (95% CI) |  |
| Overall | 214 / 15,314 | 1.40 | 264 / 15,393 | 1.72 | 0.32 (0.02, 0.61) | 256 / 15,036 | 1.70 | 242 / 15,322 | 1.58 | -0.12 (-0.41, 0.19) | 0.44 (0.00, 0.84) |
| 65-69 y | 34 / 5,513 | 0.62 | 41 / 5,370 | 0.76 | 0.15 (-0.17, 0.46) | 46 / 5,240 | 0.88 | 47 / 5,261 | 0.89 | 0.02 (-0.34, 0.36) | 0.13 (-0.34, 0.60) |
| 70-74 y | 49 / 4,240 | 1.16 | 59 / 4,265 | 1.38 | 0.23 (-0.22, 0.67) | 54 / 4,200 | 1.29 | 56 / 4,278 | 1.31 | 0.02 (-0.47, 0.51) | 0.20 (-0.46, 0.86) |
| 75-79 y | 47 / 2,735 | 1.72 | 57 / 2,912 | 1.96 | 0.24 (-0.48, 0.96) | 67 / 2,710 | 2.47 | 49 / 2,924 | 1.68 | -0.80 (-1.53, -0.06) | 1.04 (0.00, 2.07) |
| 80-84 y | 44 / 1,571 | 2.80 | 42 / 1,677 | 2.50 | -0.30 (-1.55, 0.96) | 38 / 1,596 | 2.38 | 43 / 1,608 | 2.67 | 0.29 (-0.87, 1.49) | -0.59 (-2.31, 1.13) |
| ≥85 y | 40 / 1,255 | 3.19 | 65 / 1,169 | 5.56 | 2.37 (0.62, 4.12) | 51 / 1,290 | 3.95 | 47 / 1,251 | 3.76 | -0.2 (-1.84, 1.55) | 2.57 (0.09, 4.98) |

The likelihood of being diagnosed with new AF at a primary care visit in a study practice was greater in the screening versus the control group (0.24% [92 events / 38,382 encounters] versus 0.15% [62 events / 40,003 encounters]; RD 0.08%, 95% CI 0.02–0.15). In a post-hoc analysis, the likelihood of a new AF diagnosis being made in an outpatient setting was slightly higher in screening practices although the effect estimate was imprecise (**Supplemental Table 3**).

The proportion of individuals with newly diagnosed AF who were initiated on oral anticoagulants was similar in the screening (n=194, 73.5%) and control (n=172, 70.8%) arms (RD 2.7%, 95% CI −5.5–10.4, **Supplemental Table 4**). Although use of anticoagulants was lower in the oldest patients with new AF, the majority (64.6%) of those 85 years and older in the screening group were prescribed anticoagulants.

## DISCUSSION

We conducted a pragmatic cluster randomized controlled trial of screening for AF in over 30,000 individuals from a large primary care practice network where we embedded handheld single lead ECGs into vital sign assessments. Screening using handheld ECGs was feasible with 91% of eligible individuals undergoing screening during the one-year study period. Routine screening for AF in all individuals at least 65 years of age at the time of primary care practice encounters did not result in a significant increase in newly diagnosed cases of AF at 12 months. We observed a nearly two percent increase in newly detected AF between the screening and control arms among our oldest study participants, aged 85 years and older, a result which warrants replication and further evaluation. Our difference-in-difference analysis demonstrated a small increase in new AF diagnoses in screening practices compared to the prior year, driven by individuals aged 85 years or older.

Prior studies examining point-of-care AF screening have demonstrated mixed results, and none have been conducted in the United States. Non-randomized studies of AF screening and one trial that has currently only reported screening group results have observed a positive yield in detecting undiagnosed AF.^6-12^ Three randomized controlled studies tested a point-of-care strategy. The Screening for Atrial Fibrillation in the Elderly (SAFE) study was a cluster randomized controlled trial of AF screening comprising 14,802 patients across general medical practices in the United Kingdom between 2001 to 2003.^13^ The study indicated that screening using either pulse palpation or 12-lead ECGs significantly increased detection of undiagnosed AF (1.63% versus 1.04% in the usual care arm).^13^ A cluster randomized trial conducted in general practices in the Netherlands comprising 17,107 patients compared screening with a handheld single lead ECG versus usual care between 2014−2016 did not identify a significant increase in rates of AF detection at one year (1.43% in screening versus 1.37% in usual care). Only 10% of individuals in the screening arm in this trial actually underwent screening.^14^ The Diagnosing Atrial Fibrillation (D2AF) cluster randomized controlled trial compared pulse palpation, blood pressure oscillometry, or a handheld single lead ECG in 17,976 patients at primary care practices in the Netherlands in 2018. The study observed that 1.62% of individuals in the screening arm had a new diagnosis of AF versus 1.53% in the usual care arm at one year, which was not significantly different. However, only 45% of individuals in the screening arm underwent screening.^15^ In contrast, our trial was substantially larger than these prior trials and successfully screened nearly all eligible patients.

Our results have several major implications. First, single lead ECG rhythm assessment is feasible as part of contemporary routine primary care practice. However, it is not an efficient approach to identify undiagnosed AF when applied to all individuals aged 65 years or older. Our findings are in contrast to the SAFE study,^13^ conducted nearly two decades ago and having substantial influence on current European Society of Cardiology,^16^ and the National Heart Foundation of Australia and the Cardiac Society of Australia and New Zeland clinical practice guidelines.^17^ Notably, the rate of newly diagnosed AF in the usual care arm of the SAFE study (1.04%) was substantially less than that observed in the usual care arm of our study or other more recent studies,^14,15^ suggesting the increased incidence of AF in contemporary primary care practice may mitigate the potential diagnostic benefit of single lead rhythm assessments. Nevertheless, our results suggest that screening may be effective for detecting undiagnosed AF among individuals aged at least 85 years. A previous patient-level meta-analysis of observational studies further supported an association between screening yield and age.^25^ Considering that advanced age is associated with a substantially increased risk of both AF^24^ and stroke,^26^ point-of-care screening might be best targeted at the oldest adults.

Second, routine screening for undiagnosed AF in the primary care setting using single lead ECGs increases the likelihood of diagnosis at a primary care encounter and may result in a shift in diagnosing AF to the outpatient setting. Future analyses are warranted to assess whether resource utilization and, perhaps, long-term outcomes may improve when AF management is initiated in the outpatient as compared to the emergency department or inpatient settings.

Third, point-of-care AF screening is likely to result in high rates of oral anticoagulation prescription among those with newly diagnosed AF. We observed a large proportion of individuals with newly diagnosed AF prescribed new oral anticoagulation, including those with advanced age, a subgroup in whom screening may be most effective.

The present analysis focused on the detection of new AF. Secondary analyses of anticoagulation adherence, incident stroke, and bleeding, are ongoing, although such secondary analyses of the latter two event rates will be underpowered. The 30-second single lead cardiac rhythm assessment employed in this study is an intervention that is most likely to detect persistent AF, where there is little uncertainty regarding the net benefit of anticoagulation for patients with elevated predicted stroke risk.^27^ In contrast, continuous cardiac rhythm monitoring, such as with ECG patch monitors, pick up predominantly low-burden AF^28^ for which the benefit of oral anticoagulation remains less certain.^29^ Future studies examining the effectiveness of screening for paroxysmal AF,^30,31^ and using oral anticoagulation for stroke prevention in patients with paroxysmal atrial arrhythmias are ongoing.^32,33^ We cannot exclude the possibility that small differences in temporal trends in AF incidence within the screening and control arms biased our primary overall screening effect toward the null, since a difference in difference analysis suggested a positive effect of screening on new AF diagnoses in the screening arm. Contamination in the control arm due to greater attention to heart rhythm was unlikely in our study since the proportion of newly diagnosed AF cases decreased slightly in the control arm between the prior year and study period. A high baseline level of attention to heart rhythm in our academically linked practices might partially explain the neutral result of screening but does not account for the large effect of screening in the oldest patients. Our neutral result likely reflects a low prevalence of undiagnosed persistent AF in patients 65−84 years old. Finally, the racial diversity of our patient population was limited, preventing race/ethnicity stratified results.

In conclusion, point-of-care screening for AF using a single lead handheld ECG in primary care patients is feasible at scale but does not lead to increased AF detection among all individuals aged 65 years or older. In contrast to some existing guidelines, our findings raise uncertainty about the use of single lead ECGs to opportunistically screen individuals aged at least 65 years of age for AF in primary care settings.^16,17^ Our results suggest that point-of-care screening for AF may be clinically effective among those with advanced age but this secondary result warrants further evaluation.

## Data Availability

Individual participant data will not be made available owing to the sensitive and confidential nature of the personal health information contained. Additional reasonable requests for study documentation may be made to the corresponding author.

## Acknowledgments

We thank the patients, office staff, medical assistants and clinicians from primary care practices who participated in this trial; the members of the Mass General Brigham eCare team who provided information technology support for the trial; cardiology ECG reviewers; the external advisory board for critical feedback on the study design; and additional members of our research community for their contributions to the implementation and conduct of this trial.

