## Supplemental Material for "Screening for Atrial Fibrillation in Older Adults at Primary Care Visits: the VITAL-AF Randomized Controlled Trial"

**Supplemental Methods**

*Randomization*
The constrained randomization approach accounted for factors including age, sex, comorbidity, and race distributions, AF prevalence, AF incidence, proportion of all patients anticoagulated, comorbidities, and number of patients. These features were obtained from prior year data available from a centralized data warehouse of electronic medical records (EMR) from across the Mass General Brigham hospital network[1] using validated algorithms.

*Recruitment procedures*
Patients attending a clinic randomized to the screening intervention were mailed a printed information sheet 14 days in advance of their first eligible visit to the practice describing the study as previously described.[2] Signage was posted in the clinics and information sheets were also available in the clinics for arriving patients.

*Screening procedures*
Individuals aged at least 65 years with a scheduled visit with a primary care clinician during the one-year enrollment period were identified using the electronic medical record (Epic, Verona, Wisconsin). Medical assistants were notified of eligible patients at encounters based on 1) an electronic flag in the electronic medical record at the time of the encounter, and 2) sheets containing a unique subject study identification number and barcode that were automatically printed at the time of patient arrival and registration in the clinic. Medical assistants were trained to briefly describe the study and ask patients if they would like to participate. If patients consented, the medical assistant then entered the participant's study identification number either manually or using a barcode into the AliveCor Kardia Pro (AliveCor Inc., Mountain View, CA) software running on an iPad (Apple Inc., Cupertino, CA). The medical assistant then instructed the patient to place their fingers on the handheld single lead AliveCor Kardia Mobile ECG electrodes. The automated screening results were visible to participating patients. Medical assistants were instructed to repeat the screening procedure once if the results of the initial screen were "Unclassified" or "No analysis" and to document the final screening results in the EMR (Epic, Verona, WI) along with other vital signs. Medical assistants were instructed to notify primary care clinicians verbally of a "Possible AF" result. Screening results were also viewable in the EMR by primary care clinicians at the time of the encounter. Clinicians were instructed that the AliveCor tracing results should be considered screening results only and were not diagnostic.

Two configurations of the AliveCor Kardia Mobile and iPad were used in the study, a portable handheld configuration and a mobile upright kiosk. Medical assistants were trained in a standard manner before and throughout the study regarding how to obtain an optimal tracing, including by assessing the rhythm at the end of the vital sign assessment after the heart rate had normalized, how to instruct the patient to place their fingers on the device, and how to manage the equipment.

All single lead ECG tracings were transmitted to a Web-based portal (AliveCor Kardia Pro). A team of 14 trained MGH cardiologists overread all tracings within seven days. An outcomes committee identified tracings read as AF or another prespecified potentially actionable rhythm abnormality. If the medical record indicated the primary care clinician was not aware of the abnormality, a review committee study nurse notified the clinician. Tracings were available to primary care clinicians upon request.

*AF adjudication process*
New AF was confirmed by direct search of the electronic medical record, with guidance to specifically assess prespecified elements within the chart. Newly diagnosed AF events were double-reviewed by both research nurses blinded to the other's findings. Discrepancies between nurses were resolved through consensus or by the cardiologist reviewer if uncertainty persisted. Individuals with a high probability of prevalent AF identified using a validated electronic algorithm were not manually adjudicated. In brief, this algorithm requires at least two ICD-10 codes for AF within the last three years and an anticoagulant prescription within the prior year, and has a positive predictive value of 98.4%.[3]

In a post-hoc analysis, the location of AF diagnosis was adjudicated by research nurses as either occurring in an outpatient setting (e.g. office visit, ambulatory stress test, device interrogation in an outpatient setting) or during an emergency department visit (including urgent care setting) or inpatient stay. If AF was suspected at an outpatient visit and the patient was referred directly to the emergency department for further work-up, the location of AF diagnosis was considered outpatient. All other locations were considered “other.”

**Supplemental Results**

Among eligible encounters to intervention practices in the intention-to-treat analysis during which screening was conducted, the final AliveCor algorithm result was Normal (76.8%), Possible AF (7.0%), Unclassified (12.8%), and No Analysis (3.4%) in the overall study sample, i.e., including patients with prevalent AF. Same-day 12-lead ECGs were ordered more often following encounters with Possible AF (23.2%), than following an Unclassified (9.5%), Normal (5.3%), or No Analysis (8.2%) result. Among the subset of encounters with Possible AF, the proportion of individuals who had a 12-lead ECG ordered on the same day was greater among those without prevalent AF than with prevalent AF (50.3% vs. 10.4%, $p<0.001$).

**Supplemental Table 1. Newly diagnosed atrial fibrillation in intention to treat, per protocol, and as treated analyses.**

| Analysis | Screening | | Control | | Risk difference, % (95% CI) |
|---|---|---|---|---|---|
| | No. individuals | Newly diagnosed AF, % | No. individuals | Newly diagnosed AF, % | |
| ***Individuals without prevalent AF*** | | | | | |
| Intention to treat | 15,393 | 1.72 | 15,322 | 1.59 | 0.13 (-0.16–0.42) |
| Per protocol | 14,047 | 1.77 | 14,960 | 1.60 | 0.16 (-0.14–0.48) |
| As treated | 14,409 | 1.74 | 16,306 | 1.57 | 0.17 (-0.12–0.48) |
| ***Entire study sample**** | | | | | |
| Intention to treat | 17,643 | 1.50 | 17,665 | 1.38 | 0.12 (-0.13–0.38) |
| Per protocol | 16,069 | 1.54 | 17,238 | 1.39 | 0.15 (-0.11–0.42) |
| As treated | 16,496 | 1.52 | 18,812 | 1.36 | 0.16 (-0.10–0.43) |

*Includes patients with prevalent AF.

**Supplemental Table 2. Multivariable adjusted difference in incidence of atrial fibrillation among individuals without prevalent atrial fibrillation.**

| **Model** | **Risk difference, % (95% CI)** |
|---|---|
| Unadjusted | 0.13 (-0.16–0.42) |
| Adjusted for age and sex | 0.16 (-0.12–0.45) |
| Adjusted for additional risk factors* | 0.17 (-0.12–0.46) |
| *Including age, sex, race, BMI, hypertension, coronary disease, diabetes, heart failure, indwelling cardiac devices. | |

**Supplemental Table 3. Distribution of location of new atrial fibrillation diagnoses.**

| | Screening | | Control | | Risk difference, % (95% CI) |
|---|---|---|---|---|---|
| | No. events | % events | No. events | % events | |
| Outpatient | 142 | 53.8 | 110 | 45.3 | 8.5 (-0.2–17.2) |
| Inpatient or emergency department | 118 | 44.7 | 124 | 51.0 | -6.3 (-15.0–2.4) |
| Other | 4 | 1.5 | 9 | 3.7 | 2.2 (-0.6–5.0) |

**Supplemental Table 4. New prescription of oral anticoagulation during the study period.**

| Age stratum | **Screening** No. events / No. at risk (%) | **Control** No. events / No. at risk (%) | **Risk difference, % (95% CI)** |
|---|---|---|---|
| ***Individuals with newly diagnosed AF*** | | | |
| Overall | 194 / 264 (73.5) | 172 / 243 (70.8) | 2.7 (-5.5–10.4) |
| 65-69 y | 34 / 41 (82.9) | 41 / 47 (87.2) | -4.3 (-16.9–11.8) |
| 70-74 y | 44 / 59 (74.6) | 46 / 57 (80.7) | -6.1 (-22.3–9.5) |
| 75-79 y | 40 / 57 (70.2) | 31 / 49 (63.3) | 6.9 (-6.0–27.6) |
| 80-84 y | 34 / 42 (81.0) | 29 / 43 (67.4) | 13.5 (-3.6–33.5) |
| ≥ 85 y | 42 / 65 (64.6) | 25 / 47 (53.2) | 11.4 (-2.0–29.4) |
| ***All individuals without prevalent AF*** | | | |
| Overall | 326 / 15,393 (2.12) | 312 / 15,322 (2.04) | 0.08 (-0.25–0.45) |
| 65-69 y | 70 / 5,370 (1.30) | 79 / 5,261 (1.50) | -0.20 (-0.67–0.28) |
| 70-74 y | 84 / 4,265 (1.97) | 80 / 4,278 (1.87) | 0.10 (-0.49–0.72) |
| 75-79 y | 67 / 2,912 (2.30) | 59 / 2,924 (2.02) | 0.28 (-0.45–0.98) |
| 80-84 y | 55 / 1,677 (3.28) | 45 / 1,608 (2.80) | 0.48 (-0.68–1.58) |
| ≥ 85 y | 50 / 1,169 (4.28) | 49 / 1,251 (3.92) | 0.36 (-1.24–1.98) |
